# Dimeric IgA is a specific biomarker of recent SARS-CoV-2 infection

**DOI:** 10.1101/2021.06.28.21259671

**Authors:** Heidi E Drummer, Huy Van, Ethan Klock, Shuning Zheng, Zihui Wei, Irene Boo, Rob J Center, Fan Li, Purnima Bhat, Rosemary Ffrench, Jillian SY Lau, James McMahon, Oliver Laeyendecker, Reinaldo E. Fernandez, Yukari C. Manabe, Sabra L. Klein, Thomas C. Quinn, David A. Anderson

## Abstract

Current tests for SARS-CoV-2 antibodies (IgG, IgM, IgA) cannot differentiate recent and past infections. We describe a point of care, lateral flow assay for SARS-CoV-2 dIgA based on the highly selective binding of dIgA to a chimeric form of secretory component (CSC), that distinguishes dIgA from monomeric IgA. Detection of specific dIgA uses a complex of biotinylated SARS-CoV-2 receptor binding domain and streptavidin-colloidal gold. SARS-CoV-2-specific dIgA was measured both in 112 cross-sectional samples and a longitudinal panel of 362 plasma samples from 45 patients with PCR-confirmed SARS-CoV-2 infection, and 193 discrete pre-COVID-19 or PCR-negative patient samples. The assay demonstrated 100% sensitivity from 11 days post-symptom onset, and a specificity of 98.2%. With an estimated half-life of 6.3 days, dIgA provides a unique biomarker for the detection of recent SARS-CoV-2 infections with potential to enhance diagnosis and management of COVID-19 at point-of-care.

## Introduction

Serological detection of SARS-CoV-2 infection is based on the evaluation of IgG, IgA, or IgM antibodies towards either the spike or nucleoprotein antigen with these antibody responses first detectable 5-7 days after symptom onset, with some variability ^1,2, 3,4^. A Cochrane review of 57 serological tests of 54 cohorts showed that the sensitivity at day 8-14 for IgG was 57.9-74.2% and for IgM was 45.5-70.3% with average specificity of 99.1% and 98.7%, respectively ^5^. As such, the currently available CLIA certified serological assays are considered unreliable for diagnosis of early infection. In addition, no currently available antibody assays (such as IgM or IgA) have proven reliable for the discrimination of recent from past infections, leaving a major gap in the serological tools that can be used in control of COVID-19.

As one example, rapid contact tracing has emerged as one of the pivotal public health measures to limit viral spread in the community and within health care settings. A significant gap in diagnostic testing remains for scenarios where backward contact tracing is required to identify primary source cases and their contacts who may now be SARS-CoV-2 antigen and/or RNA negative but have had recent infection, especially in healthcare settings, assisted living facilities and nursing homes, schools and high-risk workplaces. Serological approaches can be useful for backward contact tracing but this is reliant on the temporal pattern of assay sensitivity, both in the appearance of the particular antibody reactivity and in its disappearance after a predictable length of time. The low sensitivity of these serological tests, especially early in infection (<11days), limits their use in situations where every contact must be identified to track and trace recent transmission ^5^.

Respiratory infections, including SARS coronaviruses, trigger secretion of IgA, the most abundant antibody isotype produced in humans, and a major component of the mucosal immune response. IgA exists as two different isoforms, IgA1 and IgA2, with IgA2 having at least two different allotypes in the human population. In plasma, around 90% of IgA is monomeric and primarily of the IgA1 isotype, while around 10% is dimeric (dIgA), comprising two IgA monomers connected via the J chain. Although dIgA is a relatively minor component of plasma, it is the direct precursor of secretory IgA (SIgA) which is exported in large amounts on mucosal surfaces. Transport of dIgA across mucosal surfaces is performed by binding to the polymeric Ig receptor (pIgR) on the basolateral surface of epithelial cells, where it is transcytosed and the pIgR is then cleaved at the apical surface to release SIgA, representing dIgA covalently bound to secretory component (SC), the extracellular domain of pIgR. SIgA can exert multiple functions to protect against pathogens or toxins at mucosal surfaces. These include direct neutralization that blocks pathogen entry or toxin activity or binding to effector cells to activate cellular processes to enhance pathogen or toxin clearance.

All currently available tests for SARS-CoV-2 specific IgA measure total IgA, with 98.7% sensitivity (39.0-100) at day 15-21 and only 78.1% sensitivity (9.5-99.2) at day 8-14 ^3^. There is limited information on the longevity of these IgA responses, complicated by the highly variable sensitivity of IgA assays, but in a sensitive assay for spike-specific IgA in longitudinal sera the half-life was estimated to be 210 days (126-627 days) ^6^, which precludes the discrimination of recent infections. Conversely, it has previously been shown that plasma dIgA is a marker of recent infection, even when total IgA persists such as in chronic hepatitis C infections ^7^, suggesting that dIgA may provide a more reliable biomarker for recent SARS-CoV-2 infection.

In this study, we developed a novel point of care, lateral flow assay (LFA) that specifically detects dimeric IgA against the receptor binding domain of the SARS-CoV-2 spike protein in plasma and examined the temporal production of dIgA and its longevity in plasma post SAR-CoV-2 infection. Our results show that antigen-specific dIgA provides a unique biomarker of acute or recent SARS-CoV-2 infections that can be readily deployed in tests at the point of care.

## Results

### Development of lateral flow assays to measure total immunoglobulin and dIgA to SARS-CoV-2

A lateral flow test was developed to rapidly detect the presence of dIgA specific to the receptor binding domain (RBD) of SARS-CoV-2 in plasma or whole blood. The test is based on the ability of recombinant chimeric secretory protein (CSC) to specifically bind only dIgA, not monomeric IgA, IgG or IgM, as previously described ^7^. Nitrocellulose was striped with (a) CSC for the detection of dIgA, (b) RBD for the detection of total anti-RBD antibodies, and (c) anti-chicken IgY as a procedural control (Fig. 1A). Glass fiber pads were impregnated with RBD-biotin, streptavidin-Au (40nM) and chicken IgY-Au (40nM) and dried. Plasma samples (15 ul) were applied to the sample port of LFA devices followed by buffer into the buffer port. dIgA antibodies specifically binding RBD-biotin/streptavidin gold complexes are captured by the CSC stripe, while total RBD specific immunoglobulins specific to RBD are captured on the RBD stripe (Fig. 1A). Chicken IgY-Au is captured on the anti-chicken IgY line and confirms the flow of buffer through the entire nitrocellulose strip (Fig. 1B). The assay allows for visual interpretation of the presence of RBD specific dIgA and can be read using a lateral flow reader (Axxin AX-2XS, Axxin, Melbourne). A visible line corresponds to ≥400 Axxin reader units.

**Figure 1.**
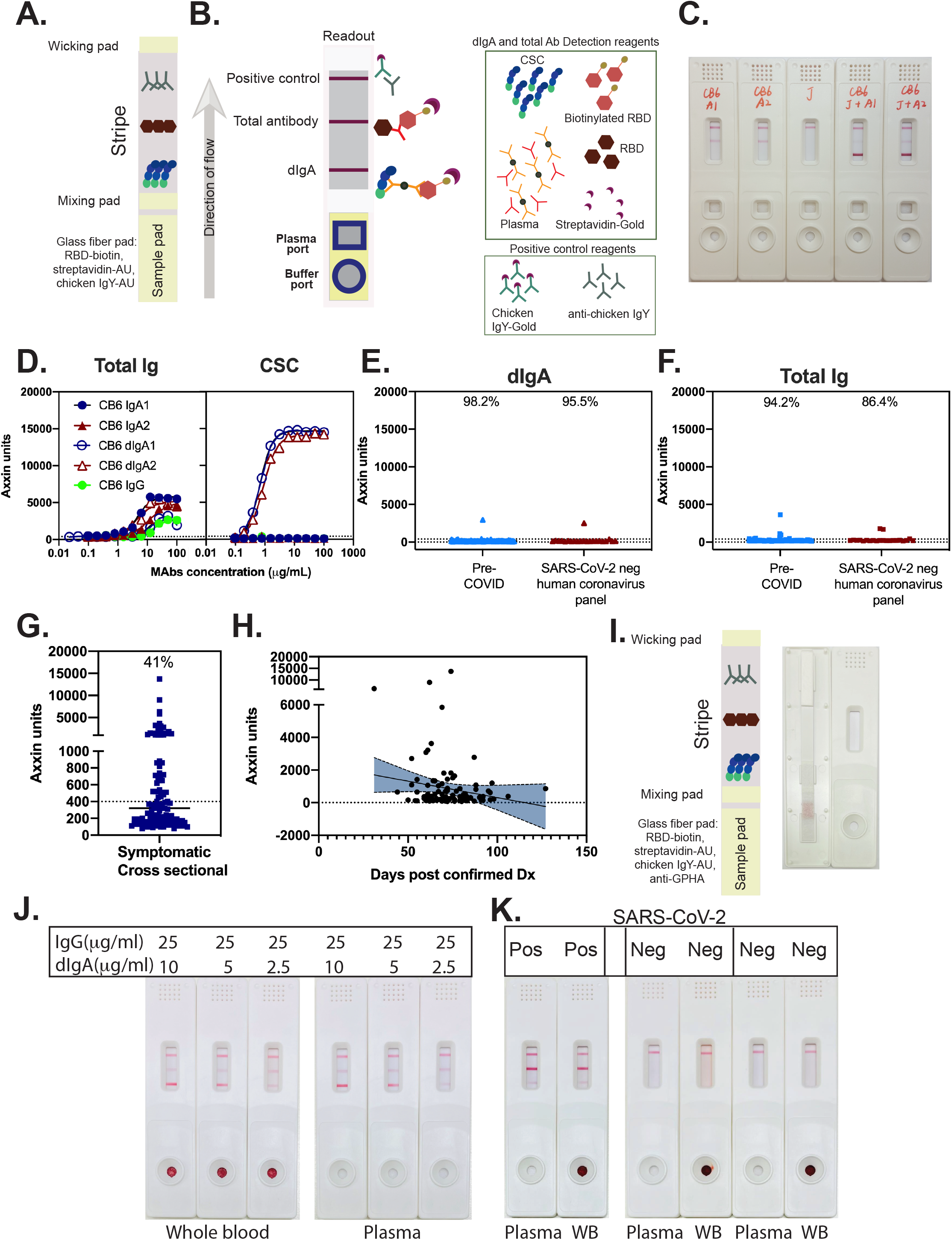
Development of a rapid lateral flow point of care assay for the detection of dIgA antibodies in plasma to SARS-CoV-2. **A**. Schematic of proteins striped onto nitrocellulose for use in LFA. **B**. Schematic of LFA test showing complexes formed at each stripe visible using 40nM gold conjugated detection reagents. **C**. Photograph of LFA test showing detection of dIgA (CB6 J+A1 and CB6 J+A2). **D**. Serial dilution of monoclonal IgA and IgG species showing dose dependent detection of immunoglobulins in the Total Ig stripe, and highly specific and sensitive detection of detection of dIgA1 and dIgA2 on the CSC stripe. **E**. Specificity of dIgA LFA. 171 pre-COVID plasmas and 22 PCR confirmed SARS-CoV-2 negative samples but with known history of non SARS-CoV-2 coronavirus infection were analysed using the LFA and measured as above. **F**. Specificity of total Ig LFA. 171 pre-COVID plasmas and 22 PCR confirmed SARS-CoV-2 negative samples but with known history of non SARS-CoV-2 coronavirus infection were analysed using the LFA and measured as above. **G**. Analysis of a 112 cross-sectional convalescent symptomatic COVID-19 subjects for the presence of dIgA. **H**. Analysis of dIgA over time in a cross sectional cohort. Linear regression analysis was performed in Prism v 9 and plotted with 95% confidence intervals. **I**. Schematic of proteins striped onto nitrocellulose for use in LFA adapted for whole blood. A anti glycophorin A sample pad was placed over the sample pad containing RBD-biotin, streptavidin-AU, and chicken IgY-AU. **J**. Examination of performance of dIgA assay for whole blood and plasma. Reconstituted whole blood or plasma was spiked with IgG and various amounts of dIgA and applied to the sample port wth buffer. **K**. Examination of performance of dIgA assay in clinical samples. Early convalescent SARS-CoV-2 (Pos) or pre-COVID-19 (Neg) plasma was tested directly (plasma) or after reconstitution as whole blood (WB).

We validated that this assay format only detects RBD-specific dIgA and not monomeric IgA by converting the Fc portion of the SARS-CoV-2 specific monoclonal antibody CB6 ^8^ to the IgA1 and IgA2 Fc sequence. Co-expression of the CB6 kappa chain with CB6-IgA1 or CB6-IgA2 heavy chain chimeras with the J chain results in dimeric IgA production; expression without the J chain results in monomeric IgA production. Immunoglobulins were purified using either ammonium sulfate precipitation and gel filtration, or via protein L affinity chromatography (Fig S1 and S2). Dimeric IgA1 and dIgA2 species eluted as 300kDa dimers in size exclusion chromatography and migrated as higher molecular weight species compared to their monomeric IgA1 and IgA2 counterparts, indicative of covalent dimerization with the J chain (Fig S2).

The tissue culture fluid of transfected cells expressing CB6-IgA1, CB6-dIgA1, CB6-IgA2, CB6-dIgA2 and J chain alone were applied to the LFA devices and showed specific detection of only dIgA1 and dIgA2 at the dIgA (CSC) test line (Fig. 1C). We next purified CB6-IgG, CB6-IgA1, CB6-dIgA1, CB6-IgA2, CB6-dIgA2 and applied serial dilutions of the proteins to the devices and measured the intensity of the lines in a lateral flow reader. The results showed highly specific detection of both dIgA1 and dIgA2 with a limit of detection of 0.123 and 0.128 µg/ml respectively, but not IgA1, IgA2 or IgG even at the highest concentrations of purified antibody (100 μg/ml) (Fig. 1D). At concentrations of dIgA that were above 3-6 μg/ml, saturation of the signal was observed; there was, however, a clear dose response within the range of 0.04-3 µg/ml. Each of IgA1, IgA2, dIgA1, dIgA2 and IgG antibodies were detectable in the total antibody (RBD) test, but with less sensitivity than for dIgA via CSC capture (limit of detection of 1.0, 1.6, 4.4, 0.7 and 5.0 µg/ml, respectively).

To examine assay specificity,171 pre-COVID-19 plasma samples, collected prior to December 2019, were examined. Of the 171 plasma samples tested, 3 were positive for SARS-CoV-2 RBD-specific dIgA by LFA providing specificity of 98.2% (Fig. 1E). Two samples were excluded from the dIgA analysis due to the presence of anti-streptavidin antibodies (not shown). For total Ig in the lateral flow test, of the 171 pre-COVID-19 samples, 10 recorded a positive result providing specificity of 94.2% (Fig. 1F).

In addition, 22 plasma samples from PCR confirmed SARS-CoV-2 negative patients with either current or previous non-SARS-CoV-2 coronavirus infection were examined to determine potential cross-reactivity with seasonal coronavirus infections. Of these 22 plasma samples, IgG reactivity towards 229E, HKU1, NL63, and OC43 was detected in 16, 21, 20, and 18 samples, respectively (Table S1). Evidence of recent or active infection with either 229E, NL63, or OC43 was detected in 1, 1, and 2 samples respectively, as determined by PCR. Using the lateral flow device, one sample returned a dIgA positive result (specificity 95.5%), while 3 total Ig positives were recorded from the non SARS-CoV-2 coronavirus panel (specificity 86.4%). None of these false positives had current PCR confirmed coronavirus infection; 2 false positive results were from individuals with HIV infection of which one had PCR confirmed parainfluenza infection.

We next examined 112 cross-sectional samples obtained from community sourced cases of PCR-confirmed SARS-CoV-2 infection for the presence of dIgA specific to RBD. Plasma samples were collected between 31-127 days post-PCR confirmation of infection and were screened for the presence of RBD-specific dIgA using the LFA. Forty plasma samples were positive for the presence of dIgA (41%) and correlated negatively with time post diagnosis, suggesting that dIgA declined rapidly during convalescence (Fig. 1G and H).

The intended use of the assay is to detect evidence of seroconversion to SARS-CoV-2 in whole blood. The assay therefore was adapted to allow the separation of red blood cells and plasma through inclusion of an anti-glycophorin A (GPHA) pad over the sample pad. Red blood cells are agglutinated via anti-GPHA and after addition of buffer, plasma flows via capillary action along the nitrocellulose membrane (Fig 1I). We compared the detection of dIgA and total Ig using whole blood spiked with a constant amount of CB6 IgG plus varying amounts of dIgA and observed a similar level of detection in both configurations (Fig 1J). Plasma from a SARS-COV-2 early convalescent subject was spiked with red blood cells to mimic whole blood and compared to plasma alone; similar levels of both dIgA and total Ig were detected. Quantitation was performed on an Axxin reader and is shown in Fig. S3. Two plasma samples collected prior to December 2019 were similarly reconstituted as whole blood or used as plasma alone and recorded negative results to both dIgA and total Ig (Fig 1K). This demonstrates proof-of-concept that the SARS-CoV-2 dIgA test can be used on whole blood to detect dIgA and total Ig to SARS-CoV-2 RBD.

### Dimeric IgA is an early marker of seroconversion to SARS-CoV-2

To investigate the timing of detectable dIgA following exposure to SARS-CoV-2, we examined dIgA levels over time in plasma samples from 45 subjects with PCR confirmed SARS-CoV-2 infection collected from -2 to 36 days after symptom onset with the majority of samples collected either 8 days before or after symptom onset (Figs S4 and S5). Samples were analysed for the presence of dIgA using the rapid LFA test and intensity measured in a lateral flow reader (Fig. 2A). In total, dIgA data were obtained for 362 plasma samples, with 25 samples omitted due to insufficient sample volume.

**Figure 2.**
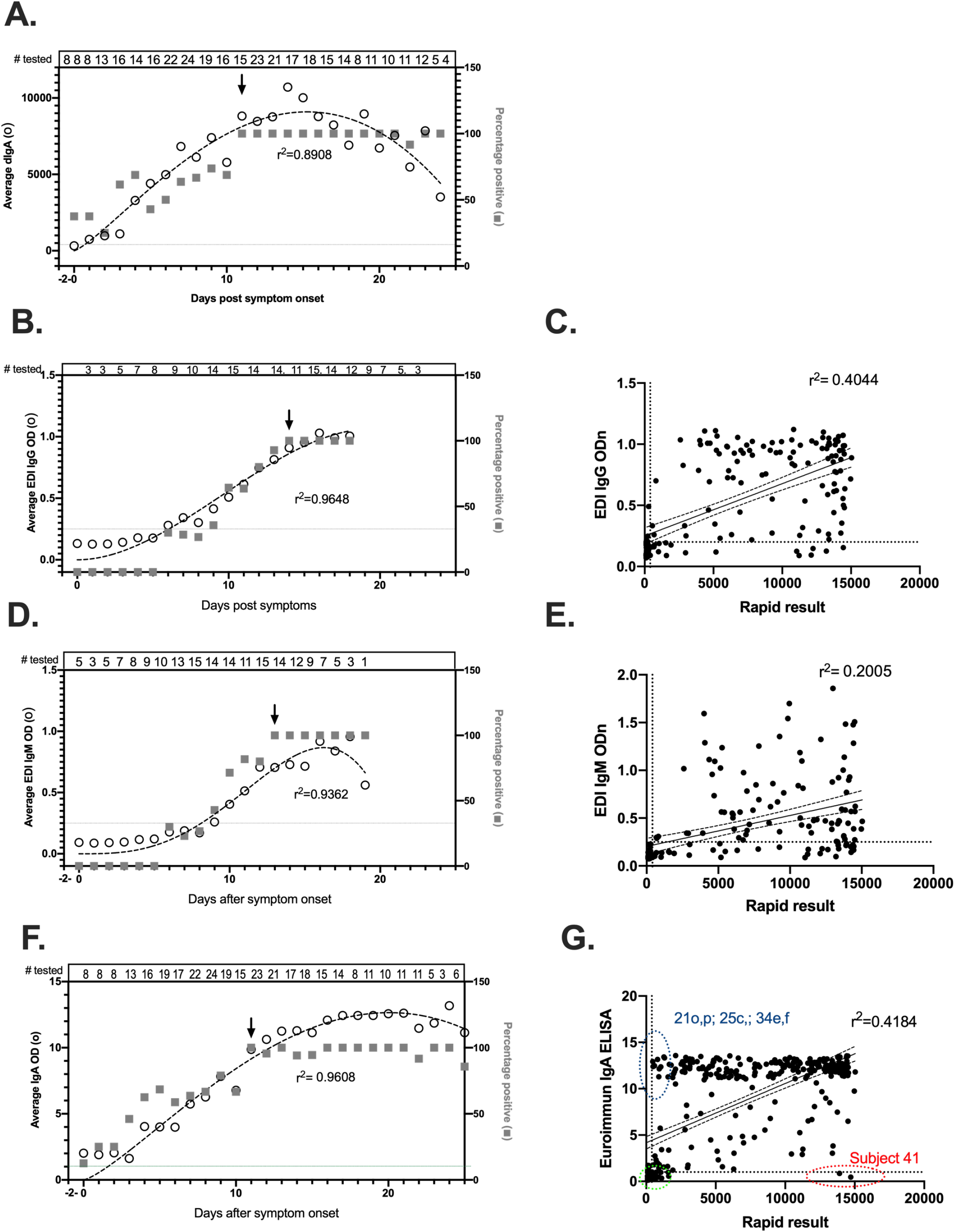
Detection of SARS-CoV-2 specific immunoglobulins in 45 subjects collected intensively post symptom onset. **A**. Number of dIgA positive samples and mean LFA read over time. Data for each time point were averaged and plotted against days post-symptom onset for days up to day 25. Day 0 includes samples from between days -2-0. A beta decay curve fitted to the data using Prism v9. Arrow indicates when 100% of samples tested positive for each assay type. **B**. Number of IgG positive samples using the EDI IgG test and mean OD over time. A beta decay curve fitted to the data using Prism v9. Arrow indicates when 100% of samples tested positive for each assay type. **C**. Correlation of IgG and dIgA results. A non-parametric Pearson correlation analysis was performed in prism v9.02. **D**. Number of IgM positive samples using the EDI IgM test and mean OD over time. Arrow indicates when 100% of samples tested positive for each assay type. **E**. Correlation of IgM and dIgA results. A non-parametric Pearson correlation analysis was performed in prism v9.02. **F**. Number of IgA positive samples using the EUROIMMUN IgA test and mean OD over time. Arrow indicates when 100% of samples tested positive for each assay type. **G**. Correlation of IgA and dIgA results. A non-parametric Pearson correlation analysis was performed in prism v9.02. The number of samples testing positive at each day post symptom onset is shown on the right axis and the number of samples tested is shown above each graph.

Results indicate that between 0 and 10 days after COVID-19 symptom onset, there was a steady increase in the levels of dIgA as well as the number of subjects who tested positive (>400 Axxin units) for dIgA (Fig. 2A and Fig S4), with 100% of subjects having detectable dIgA by day 11 post-symptom onset, with an average of 8817 units. Peak levels of dIgA were detected between day 14 and 15 after symptom onset. Overall, dIgA was detected in 49% of people between 0-5 days, 65% between days 6-10, and 100% from day 11 onwards post-symptom onset (Table 1). While the number of subjects testing positive was maintained over 25 days, the average level of dIgA steadily declined from day 15 onwards.

**Table 1.** Detection of SARS-CoV-2 specific antibody isotypes days post PCR diagnosis.

| Antibody type<sup>1</sup> | Days post PCR +ve (%)<sup>2</sup> |  |  | Calculated days from PCR diagnosis to earliest 100% positive<sup>3</sup> |
| --- | --- | --- | --- | --- |
|  | 0-5 | 6-10 | 11-15 |  |
| dIgA | 49 | 65 | 100 | 11 |
| IgM | 0 | 39 | 92 | 13 |
| IgG | 0 | 34 | 67 | 14 |
| IgA | 47.2 | 69 | 96.8 | 11 |

1 Plasma samples were analysed in the dIgA LFA (dIgA), EDI IgM ELISA (IgM), EDI IgG ELISA (IgG) and EUROIMMUN IgA ELISA (IgA).
2 The percentage of samples tested that recorded a positive result
3 Number of days post symptom onset before 100% of samples tested positive.

We analysed the same subjects for the presence of IgG and IgM specific to nucleocapsid (N) antigen. The test utilised was reported to be able to detect IgM in 5.9% of people ≤ 5 days after symptom onset, 37.1% for IgM between 5-10 days, 76.4% for IgM 10– 15 days, and 94.4% for IgM after days 15-22. The reported “false” positivity rates were 0.5% for IgM in healthy blood donors and 1.6% for IgM in ICU patients^9^. Likewise, for IgG, 37.1%, 82.4% and 100% of samples were reported to test positive between days 5-10, 10-15 and 15-22, respectively, with a false positive rate of 1% and 1.2% for healthy blood donors and ICU patients, respectively^9^. In this study, none of the samples tested were positive for SARS-CoV-2 anti-N IgG or IgM in the first 5 days (Fig. 2B and 2D and Fig S4). Between days 6-13, there was a steady increase in the number of samples testing positive, and by day 13 (IgM) and day 14 (IgG), all samples tested were positive for antibody against SARS-CoV-2 N protein. Overall, between days 0-5, 6-10 and 11-15, 0%, 39% and 92% of samples tested positive for IgM and 0%, 34% and 67% tested positive for IgG, respectively (Table 1). While dIgA and IgG were moderately correlated r^2^=0.4044 (p<0.0001) (Fig. 2C), dIgA and IgM were relatively weakly correlated, r^2^=0.2005 (p<0.0001) (Fig 2E).

To compare detection of dIgA with total IgA, we analysed the same subjects for the presence of IgA using a commercially available test that detects IgA against the S1 domain of SARS-CoV-2 (EUROIMMUN IgA). The reported specificity of the EUROIMMUN IgA assay was 88.4% and sensitivity was 82.9%^10^. Both dIgA and anti-S1 IgA show a similar trajectory early in infection up to day 20 (Fig 2F and Fig S4). After day 15, the levels of dIgA declined, while the levels of anti-S1 IgA were maintained. Overall, between days 0-5, 6-10, and 11-15, 47.2%, 69 %, and 96.8% of samples, respectively, were positive for IgA (Table 1). Anti-S1 IgA and dIgA were correlated (Fig 2G) (r^2^=0.4184, p<0.0001). There were, however, exceptions, including one individual who had particularly high levels of dIgA but was negative for anti-S1 IgA activity (Fig 2G). There also was a cluster of samples that were high for anti-S1 IgA but low for dIgA. The EUROIMMUN IgA assay also appeared to reach a maximal level of detection as a large number of data points cluster at a sample/cutoff ratio of ∼12-13 whereas dIgA measured in the dIgA LFA assay appears to have a wider dynamic range. Taken together, these data suggest that dIgA may be an acute phase serological marker of SARS-CoV-2 infection, that dIgA is generated earlier than both IgM and IgG and follows a similar trajectory to IgA early in infection. Unlike total IgA, dIgA levels decline more rapidly in plasma beyond 15 days after symptom onset.

### Half-life analysis of dIgA

The ability to detect dIgA earlier than either IgG or IgM after SARS-CoV-2 infection suggests it may have complementary utility to antigen or PCR-based detection of active SARS-CoV-2 where suspected cases return a negative result, in order to increase diagnostic sensitivity and for backward contact tracing to identify recently infected antigen- or PCR-negative individuals. We therefore determined the half-life of dIgA to determine how long dIgA persists in plasma to provide a window of infection recency using a well characterised seroconversion panel. In addition, we analysed the same samples for the presence of anti-S1 IgG and IgA using the EUROIMMUN IgG and IgA ELISA. Five of the 6 subjects in this panel were dIgA positive and were used to calculate the half-life (Fig. 3). A one-phase decay equation was used to calculate the half-life of dIgA observed in the rapid LFA with an average r^2^ of 0.9862. The mean interval between first and last sample included for calculation was 62 days. The half-life of dIgA was 6.3 days with lower and upper 95% confidence intervals of 3.6 and 9.6 days, respectively.

**Figure 3.**
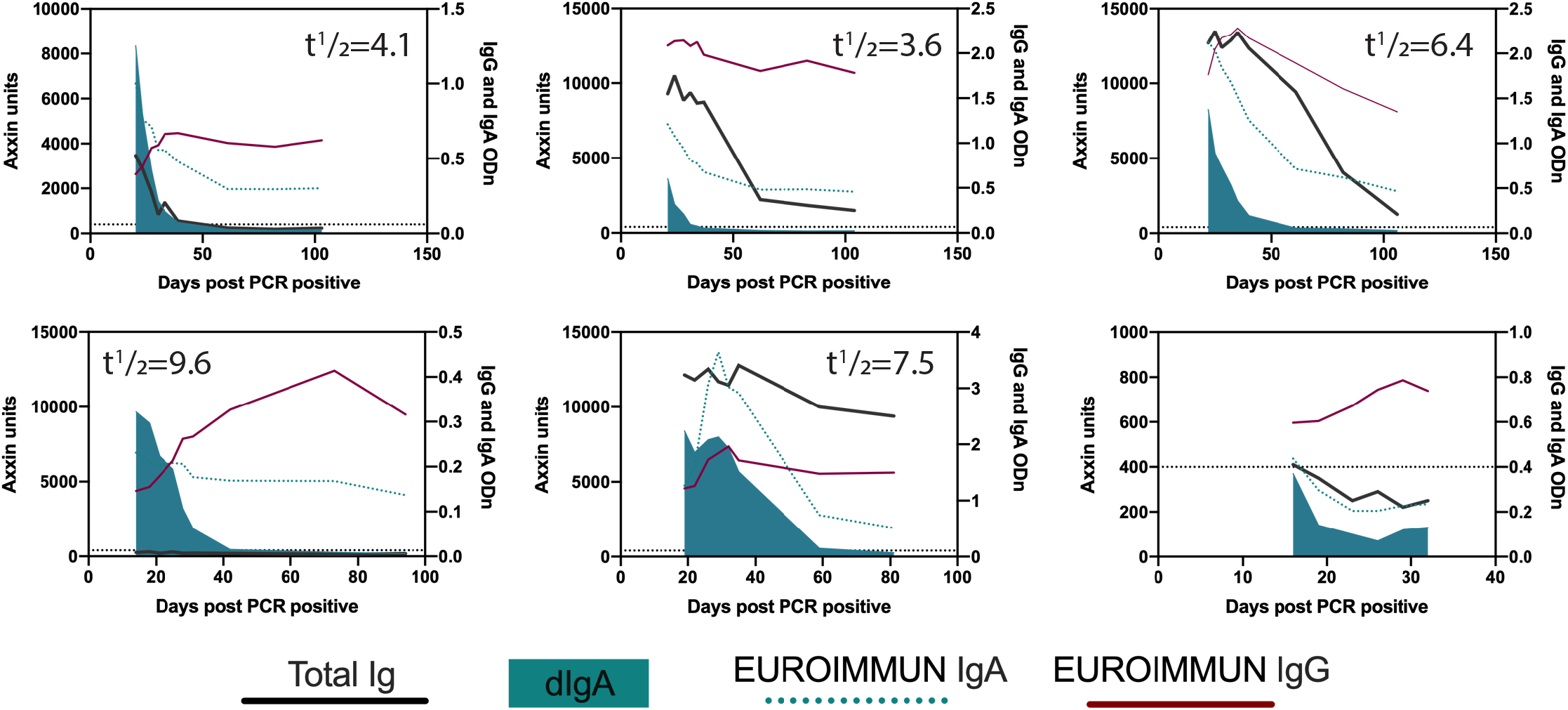
Half-life of dIgA in longitudinal samples. dIgA and Total Ig levels were measured using the rapid LFA and data fitted using a one-phase decay equation (Prism v 9.02). The dIgA half-life is shown for each subject. The same samples were analysed for the presence of IgG and IgA using the EUROIMMUN ELISA and are plotted on the right y-axis.

A similar analysis performed for the total immunoglobulin line of the LFA test revealed that 5 of these 6 individuals had a detectable total immunoglobulin positive result, of which 1 became negative within the observation period at day 19 post PCR. The levels of total Ig in the remaining 7 individuals declined at a slower rate over the sampling period compared to dIgA and a broad range of half-life values was calculated, with low confidence (average r^2^=0.88) ranging from 6.4 to 207,143 days (Fig 3). By contrast, the anti-S1 IgG levels in these same samples remained relatively steady and did not fit the one phase decay nor a continuous decay equation. Anti-S1 IgA consistently displayed a 2-3-fold longer half-life relative to dIgA for each of the subjects, with a mean half-life of 13.88 days (r^2^=0.9668) using a one-phase decay equation.

## Discussion

In this report, we describe antigen-specific dIgA as a potential new biomarker of recently acquired SARS-CoV-2 infection, deployed in a rapid point of care lateral flow assay. The assay is based on the ability of a chimeric (rabbit/human) form of the polymeric Ig receptor, CSC, to bind with high specificity to dimeric IgA^7^, thereby immobilising complexes of antigen-specific dIgA and SARS-CoV-2 RBD antigen-colloidal gold. The ability of CSC to bind dimeric IgA exclusively was confirmed using purified dimeric and monomeric IgA specific to the SARS-CoV-2 RBD, with no reactivity observed towards monomeric IgA or IgG forms of the same immunoglobulin when present in ≥200-fold excess of the limit of detection for dIgA. Application of this assay to a large panel of pre-COVID plasma and a panel of samples from confirmed non-COVID-19 coronavirus infection showed assay specificity of 98.5%. The prototype assay can be performed on 15ul of plasma or 30µl of whole blood in under 30 minutes and provides a rapid assessment of serological activity towards SARS-CoV-2.

Using a longitudinal panel of 362 samples sourced from -2 to 36 days after symptom onset from 45 subjects, we determined that between days 0-5, dIgA was detectable in 49% of samples, reaching a peak of 100% detection by day 11. By contrast, IgM, usually considered an early marker of seroconversion, was absent before day 5, and only detected in 39% of subjects between days 6-10, reaching a maximum of 92% detection between days 11-15. This was not dissimilar to IgG with 34% detection between days 6-10 and 67% between days 11-15. Comparison with IgA responses in the same subjects revealed that whilst the trajectory was similar, IgA appeared to be more stable in the plasma and reached assay saturation in many subjects. By contrast, the dIgA LFA responses appear spread over a wide dynamic range and were consistently recorded after 11 days post symptom onset with a measurable decline in the level of reactivity beyond 20 days. One subject in particular did not generate a detectable IgA response at either time point tested but was strongly positive for dIgA.

Dimeric IgA is actively transported across mucosal surfaces by binding to pIgR on the basolateral surface of epithelial cells. After transcytosis, pIgR is cleaved at the apical surface to release SIgA covalently bound to SC. This process actively depletes dIgA from plasma over time, concentrating dIgA instead to the mucosa where it protects against infection by pathogens entering via mucosal surfaces. Our analysis of the large longitudinal cohort revealed that the levels of dIgA appear to begin to decline after approximately day 20. This cohort extended to at most day 35 post symptom onset, with many patients exhibiting a plateau of dIgA reactivity through the period of sampling (Figure S4). To examine the half-life of dIgA, we analysed two extended longitudinal panels where there were at least four data points showing a decline in dIgA levels. Six subjects were included in this analysis and revealed that the half-life of RBD-specific dIgA in serum was 6.3 days. A recent study using 844 samples collected from longitudinal samples from 151 RT-PCR positive convalescent SARS-CoV-2 patients showed that the half-life of RBD specific IgG antibodies was 62 days ^11^, while a separate study of 51 patients reported even longer half-lives of spike-specific IgG in paired serum samples of 103 days and for IgA 210 days^6^. While the observed short half-life of SARS-CoV-2 specific dIgA precludes its use for measuring past infections beyond a few months, when combined with highly sensitive detection in the early phase after infection (peaking at 100% between 11-35 days) this instead provides a unique serological biomarker for recent or active infection.

Serology for many viral infections relies on the detection of IgM as a marker of acute or current infection, with IgG as a marker of past infection, and the early part of the COVID-19 pandemic saw a profusion of commercially available laboratory and POC tests designed for the measurement of SARS-CoV-2 specific IgG and IgM. However, IgM has many limitations in serology especially with low-prevalence infections ^12^, and none of these COVID-19 IgG/IgM tests have proven to be useful for determining the timing of recent or past infections in the absence of direct virus detection, severely limiting their utility in public health control measures.

Our results suggest that SARS-CoV-2 specific dIgA may provide this missing tool for COVID-19 serology. Rapid antigen detection tests have received emergency authorization use under the FDA and have shown high specificity (>99%). However, the sensitivity of such tests is highly variable in symptomatic patients (63.7-79%) and have highest sensitivity (94.5%) where there is a high viral load (Ct values ≤25 or >10^6^ genomic virus copies/mL) early in infection (<7 days) ^13^. The dIgA test performs at its best in the 2^nd^ and 3^rd^ week post symptom onset and a positive result by itself would indicate recent infection. The use of a rapid antigen test and the dIgA POC test in combination could allow rapid mass screening of individuals to identify active and recent past infections within at least the past 21 days, which would greatly enhance contact screening. For example, the dIgA LFA test could be used in backward contact tracing, including well-recognised super-spreader events. In these situations, where one or more individuals later become symptomatic and return a positive nucleic acid or antigen test result, there is often a missing link to the original source of infection because antigen and nucleic acid tests are likely to have fallen below the limit of detection by the time contact tracing is initiated. Wide-spread dIgA testing could be deployed to determine those who have evidence of recent SARS-CoV-2 infection to track chains of transmission, identify potential common sources of infection and their onward contacts, and reducing the number of people who need to isolate in order to reduce forward transmission of the virus.

Our study used samples collected from individuals with symptomatic infection. We have not examined the assay sensitivity in people with asymptomatic infection. Studies have shown that the levels of IgG in asymptomatically infected people are lower in the acute phase and decline faster than those showing symptoms ^14^. It is possible that the same is true for dIgA and further analysis of plasma from verified asymptomatically infected people is required to examine this more closely. Our total immunoglobulin test line showed relatively poor specificity of 93.3%. The reasons for this may relate to the use of the isolated receptor binding domain in a double sandwich allowing low affinity interactions to be maintained, particularly relevant for IgM antibodies. Improvements to the total Ig line are required before this aspect of the test could be deployed in the field.

While the role of IgA in neutralization of SARS-CoV-2 virus and its appearance after infection have been documented, this is the first study to document the appearance of dIgA in the plasma of SARS-CoV-2 subjects. Studies report the appearance of IgA as early as 2 days after symptom onset, earlier than IgG or IgM, detected for both nucleocapsid and RBD. This is consistent with our observations here that dIgA can be detected in 49% of subjects testing PCR positive up to 5 days after symptom onset, and 100% at day 11^15^. Serum neutralization titres also correlate more strongly with IgA specific to RBD than anti-RBD IgG titres and purified monomeric IgA is also more potent than IgG at mediating neutralization of SARS-CoV-2^15^. Examination of saliva and bronchoalveolar lavage reveal that IgA dominates the neutralization activity, with dimeric IgA detected in bronchoalveolar lavage fluid^15^. Isolation of B cell clones expressing IgA towards the SARS-CoV-2 spike protein and the expression in monomeric or dimeric form reveals that dimeric IgA is 3.8-113-fold times more potent than the corresponding IgA monomers at neutralizing pseudoviruses and 15 -fold higher for authentic SARS-CoV-2 virions^16^. This suggest that dIgA plays an essential role in mucosal protection against SARS-COV-2 and may indeed be a potent contributor to antibody neutralization activity, and our development of dIgA-specific tests should contribute to improved understanding of these factors. The results presented here suggest that dIgA provides a novel and readily detected biomarker of recent SARS-CoV-2 infections, with particular utility for contact tracing and potential for improved detection of incident infections when used in conjunction with rapid antigen tests.

## Materials and Methods

### Cells and cell lines

Expi293F cells were maintained in suspension culture in Expi293 expression medium (ThermoFisher Scientific) at 37°C and 8% CO_2_. Plasmids encoding proteins of interest were transfected into cells using the ExpiFectamine 293 reagent according to the manufacturer’s instructions (ThermoFisher Scientific). In order to biotinylate proteins with a C-terminal Avitag (GLNDIFEAQKIEWHE), Expi293F cells stably transfected with a plasmid directing expression of the BirA biotin ligase (ExpiBirA) were used (a kind gift of Dr Bruce Wines). For in situ biotinylation, media was supplemented with a final concentration of 50 μM D biotin.

#### Vectors

The receptor binding domain (RBD) of the SARS CoV-2 spike protein (amino acids 332-532) and the chimeric secretory component (CSC) of the polymeric immunoglobulin receptor (650 amino acids protein synthesized by GenScript) were synthesized by ThermoFisher Scientific and subcloned into pcDNA3-based vectors under the control of the CMV promoter^7^. Both proteins encode a C-terminal 6 histidine tag to enable purification by immobilized metal affinity chromatography (IMAC). In addition, the Avitag sequence was appended onto the C-terminus of RBD to enable site-specific biotinylation in ExpiBirA cells.

#### Protein Expression and purification

After transfection with RBD-encoding plasmid Expi293F or Expi293F BirA cells were incubated at 34°C whereas Expi293F cells transfected with CSC-encoding plasmid were incubated at 37°C. Four days after transfection tissue culture supernatants were clarified and target proteins were purified by IMAC using Talon metal affinity resin (Clontech Laboratories) following the manufacturer’s recommendations. The eluted proteins were subject to gel filtration using a Superdex 200 16/600 column (GE Healthcare) with PBS as the liquid phase. Fractions corresponding to monomeric RBD or dimeric CSC were pooled and concentrated in Amicon Ultra 10 (RBD) or 30kDa (CSC) devices (Merck) prior to use.

#### ELISA

Plasma samples were tested in ELISA based assays to determine levels of IgA (EUROIMMUN AG), IgG (EUROIMMUN AG and EDI Inc), and IgM (EDI Inc) according to the manufacturer’s instructions.

#### Antibodies and proteins

Recombinant synthetic (GeneArt) chimeric IgA1 and IgA2 heavy chain sequences were constructed by joining the CB6 variable domain (amino acids 1-137) to the CH1 domain (amino acids 161-516) of IgA1 (J00220) or the CH1 domain (amino acids 161-501) of IgA2 (J00221) via a BspE1 restriction site and cloning into a pcDNA3 vector with an N-terminal TPA leader sequence. For the expression of the J chain the mature protein sequence (DQ884395) amino acids 22-175 was synthesized (GeneArt) and cloned in frame with the tissue plasminogen activator (tpa) leader sequence into pcDNA3 based expression vector (Fig S1). Recombinant chimeric IgA1 and IgA2 were expressed in 293 Expi cells by co-transfection of the CB6 kappa chain and CB6-IgA1 or CB6-IgA2 heavy chain sequence. Dimeric IgA1 and IgA2 were expressed by co-transfecting CB6 kappa light chain and CB6-IgA1 or CB6-IgA2 heavy chain sequence and pcDNA3-J using equivalent amounts of DNA. After 4 days, supernatant fluid was collected, clarified and either used directly, or purified using 45% ammonium sulfate and size exclusion chromatography on a Superose 6 Increase 10/60 GL column (AKTAPure). Proteins were analyzed on SDS-PAGE and visualized with Coomassie blue.

The construction and expression of recombinant synthetic CSC protein has been previously ^7^. The following reagents were sourced from commercial suppliers: Streptavidin-gold (40nM) was purchased (Abcam cat# ab186864), Chicken IgY gold conjugate 40nm (BBI cat# Cont.Chick), SARS-CoV-2 RBD (GenScript Cat# Z03483-1), 5D protein stabilizer (Abacus cat# 5D82411B)

#### Lateral flow assay

The device cassette consists of a plastic housing (Nanjing BioPoint Diagnostics, PR China) with loading wells and a window to read results. Within the cassette is a nitrocellulose membrane strip (Sartorius, Germany), with a conjugate pad containing biotinylated RBD protein complexed with streptavidin-colloidal gold and colloidal gold-conjugated chicken IgY as a procedural control. RBD-biotin (4ug/mL) was mixed with streptavidin gold conjugate (OD 1.5) for 30 min at room temperature. Chicken IgY gold conjugate (OD 1) and drying buffer (5mM HEPES, 0.17% PEG, 1mM EDTA, 0.5% BSA, 10% trehalose, 0.12% Tween) were added to the mixture and dispensed onto conjugate pads, frozen at -80°C for 30 min and vacuum dried for 1 h at room temperature. Nitrocellulose membranes (Sartorius, Germany) were striped with proteins in PBS (pH 7.4) to give two test lines, with test line 1 comprising 0.4 µl/cm of CSC (1 mg/ml) to capture dIgA, and test line 2 comprising 0.4 µl/cm of RBD (0.5 mg/ml) to capture total anti-RBD antibody, and a third procedural control line comprising 0.4 µl/cm of rabbit anti-chicken IgY (0.1 mg/ml) and dried at room temperature. Conjugate pads and nitrocellulose membranes were laminated together with absorbent pads at the distal end and cut into 5 mm wide strips before placing into cassettes.

The test specimen (15µl plasma) is dispensed into the sample well A (plasma port) of the test cassette, rehydrating the gold conjugates, and four drops of running buffer (phosphate buffered saline pH 7.4, 0.5% Tween20, 0.05% Sodium azide) are added to well B (buffer port) of the test cassette, initiating sample flow by capillary action. The test is read after 30 minutes. For whole blood tests, in the absence of fresh patient samples we used plasma that was reconstituted by addition of 15µl plasma to 15µl packed red blood cells. The device was modified by addition of a glass fibre soaked in anti-glycophorin A over the sample pad, and both sample and buffer were added to well B (buffer port) while the plasma port (well A) was not used. Whole blood (30 µl) or plasma (15 µl) was added to the buffer port with the addition of four drops of running buffer. The result was read after 30 minutes.

#### Statistical methods

Data was analysed in Prism v9.02. Normality was determined using the D’Agostino & Pearson test. Data were correlated using the nonparametric Spearman test, two-tailed p value with 95% confidence intervals. A simple linear regression was used to calculate lines of best fit and r2. To fit appearance of IgG/A/M over time, a beta growth then decay curve was fitted to the data. To determine the half-life of dIgA, a one-phase decay equation was used with no constraints.

### Ethics

This study was conducted in concordance with the principles of the Declaration of Helsinki of the World Medical Association and approved by local human research ethics committees. Ethical approval was granted for this work by Alfred HREC (296/20: Development of serological tests for COVID-19) and approved by The Johns Hopkins University School of Medicine Institutional Review Board (IRB00247886, IRB00250798 and IRB00091667). Pre-pandemic negative control sera were provided by Australian Red Cross Lifeblood (Material supply agreement 20-08 Vic-05). All samples were de-identified prior to testing. This report includes an analysis of stored samples and data from those studies. No additional samples were collected for the current study.

## Data Availability

All data is presented in the manuscript. Data files can be obtained from Authors upon request.

## Author contributions

HED, HV, EK, SZ, ZW, IB, RJC, FL, PB, RF, JSYL, JMcM, OL, REF, YCM, SLK, TCQ, DAA, performed experiments, provided samples or reagents and/or directed experimental work. HED and DAA conceived the study, analysed the data, co-wrote the paper and co-led the project.

