## Supplemental files for "Dimeric IgA is a specific biomarker of recent SARS-CoV-2 infection"

**A.**

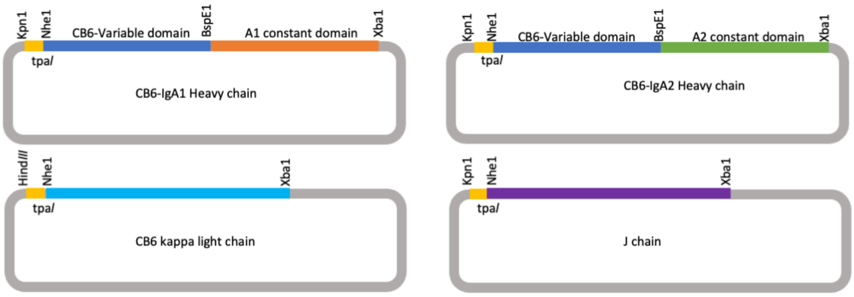


**B.**

| Protein | CB6 kappa chain | CB6 Heavy chain | J chain |
|---|---|---|---|
| CB6-IgA1 | 15 µg | 15 µg IgA1 | - |
| CB6-dIgA1 | 10 µg | 10 µg IgA1 | 10 µg |
| CB6-IgA2 | 15 µg | 15 µg IgA2 | - |
| CB6-dIgA2 | 10 µg | 10 µg IgA2 | 10 µg |
| J chain | - | - | 30 µg |

**C.**

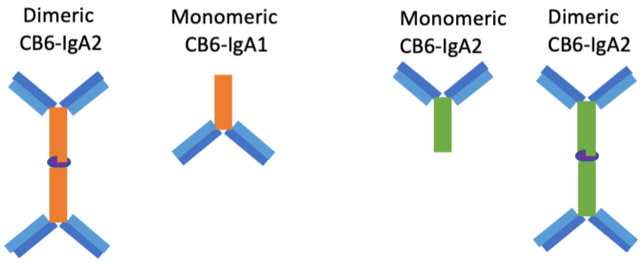


**Figure S1.** A. Schematic of vectors encoding chimerized DNA sequences for the expression of CB6-IgA1, CB6-IgA2, CB6-dIgA1 and CB6-dIgA2 immunoglobulins. B. Transfection conditions for expression in 293 Expi cells at 30 ml scale. C. Schematic of IgA species used to validate lateral flow test.

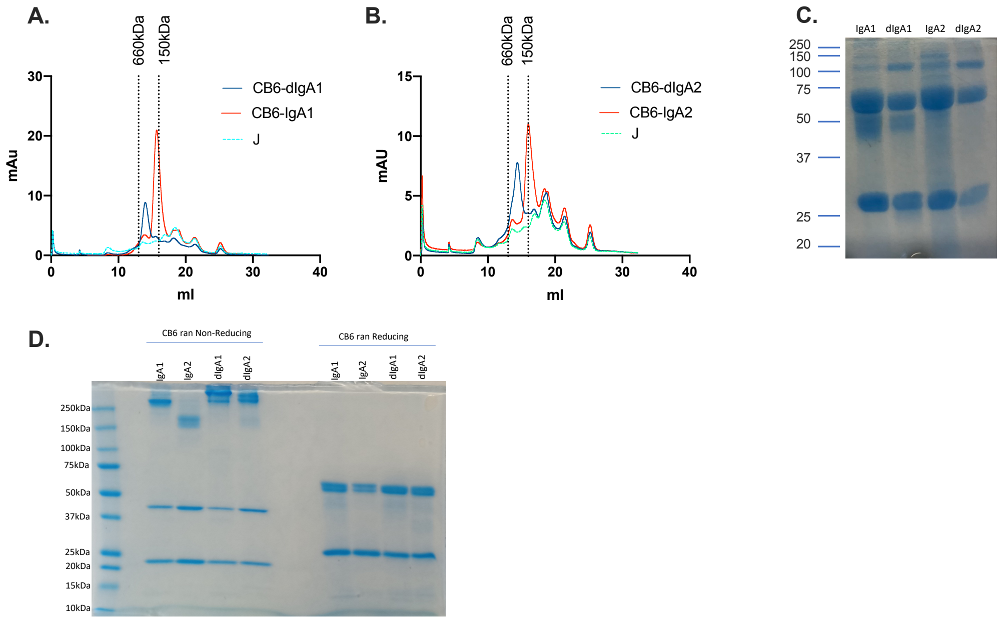


**Figure S2.** Purification of IgA and dIgA species. Ammonium sulfate precipitated CB6-dIgA1 and monomeric CB6-IgA1 (A) and CB6-dIgA2 and monomeric CB6-IgA2 (B) were buffer exchanged by dialysis into PBS and proteins purified by size exclusion chromatography on a Superose 6 10/30GL column. Peaks corresponding to the predicted size of dIgA1/2 and monomeric IgA1/2 were collected, concentrated and examined in SDS-PAGE (C). Protein L purified IgA species were affinity purified using Protein L and examined by 8-15% SDS/PAGE either non-reduced or reduced.

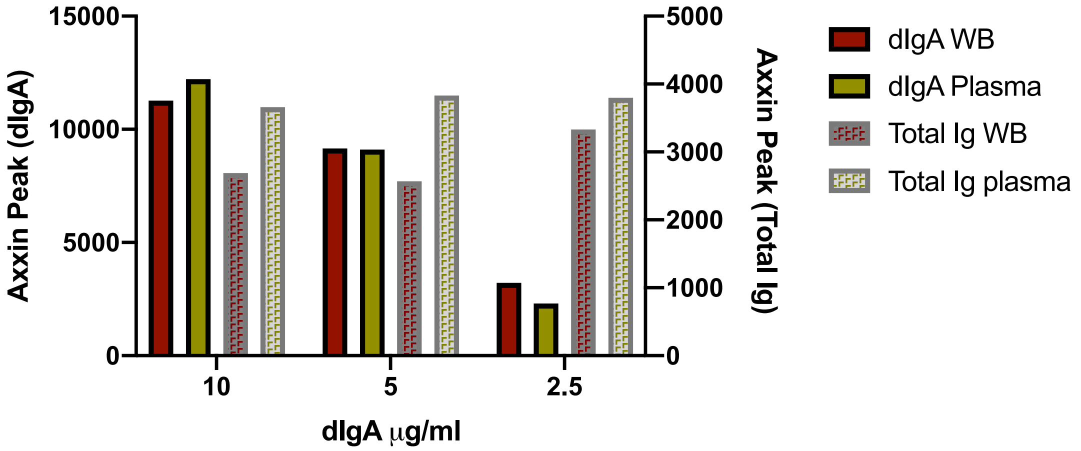


**Figure S3.** Quantitation of dIgA and total Ig levels in whole blood or plasma using the dimeric IgA LFA. Samples shown in Figure 1J were quantitated in an Axxin reader.

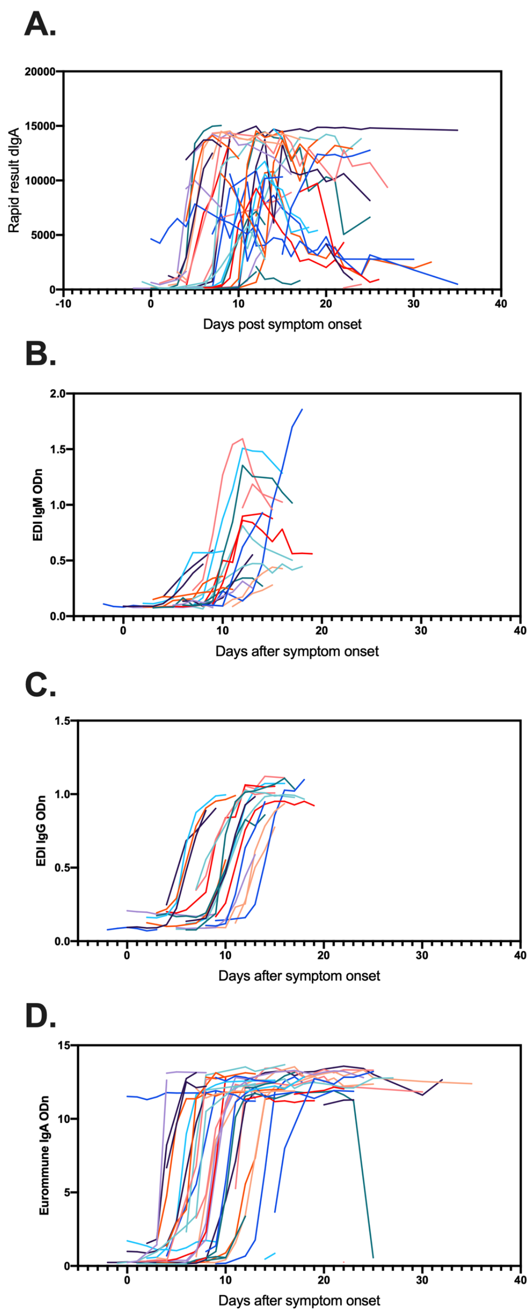


**Figure S4.** Analysis of dIgA in lateral flow assay (A), IgM using EDI ELISA (B), IgG using EDI ELISA (C) and IgA using EUROIMMUN ELISA (D). Plasma samples obtained at various times post symptom onset from 45 subjects were examined for the presence of each antibody type.

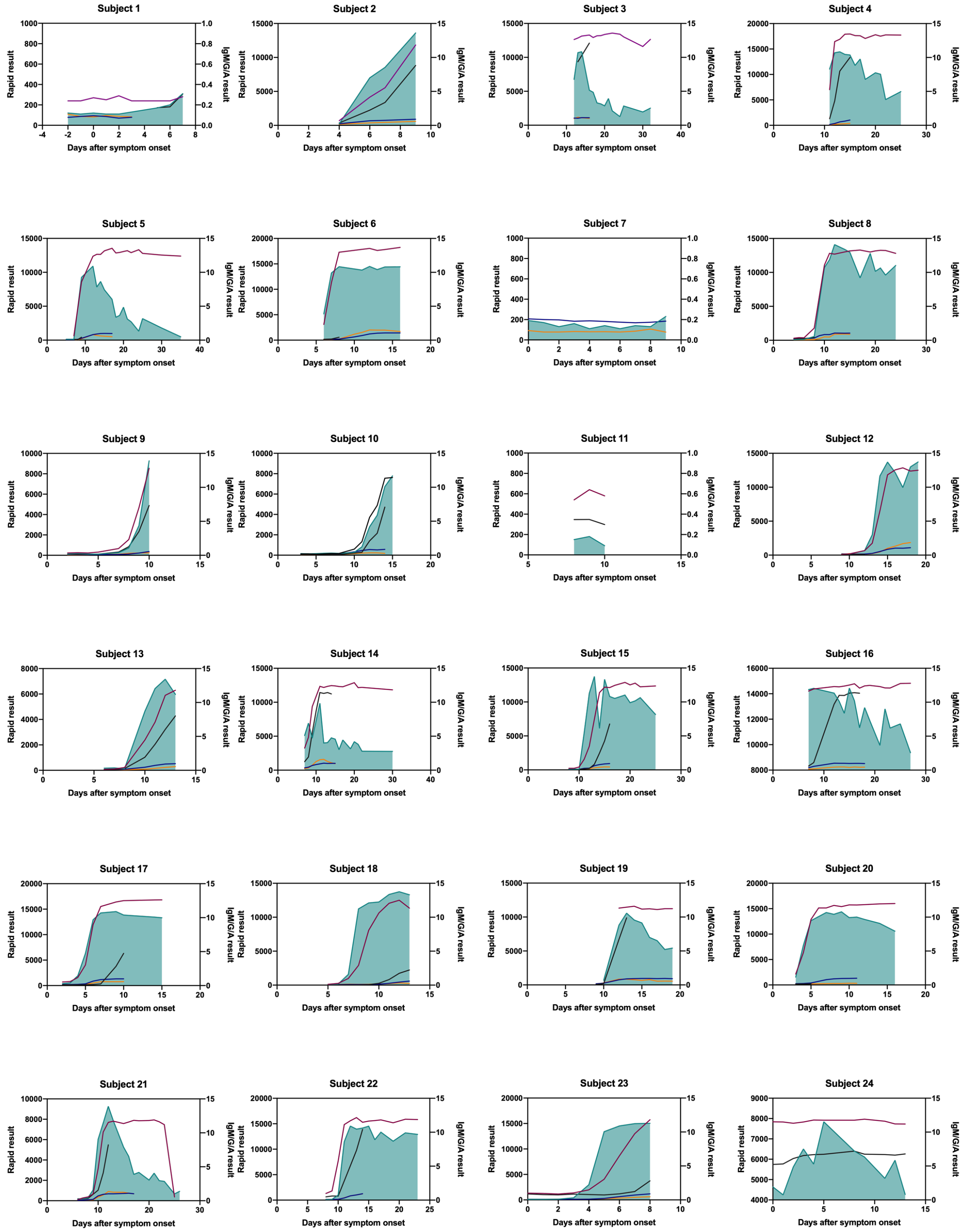
Subject 1
Subject 2
Subject 3
Subject 4
Subject 5
Subject 6
Subject 7
Subject 8
Subject 9
Subject 10
Subject 11
Subject 12
Subject 13
Subject 14
Subject 15
Subject 16
Subject 17
Subject 18
Subject 19
Subject 20
Subject 21
Subject 22
Subject 23
Subject 24
Rapid result
IgM/G/A result
Days after symptom onset

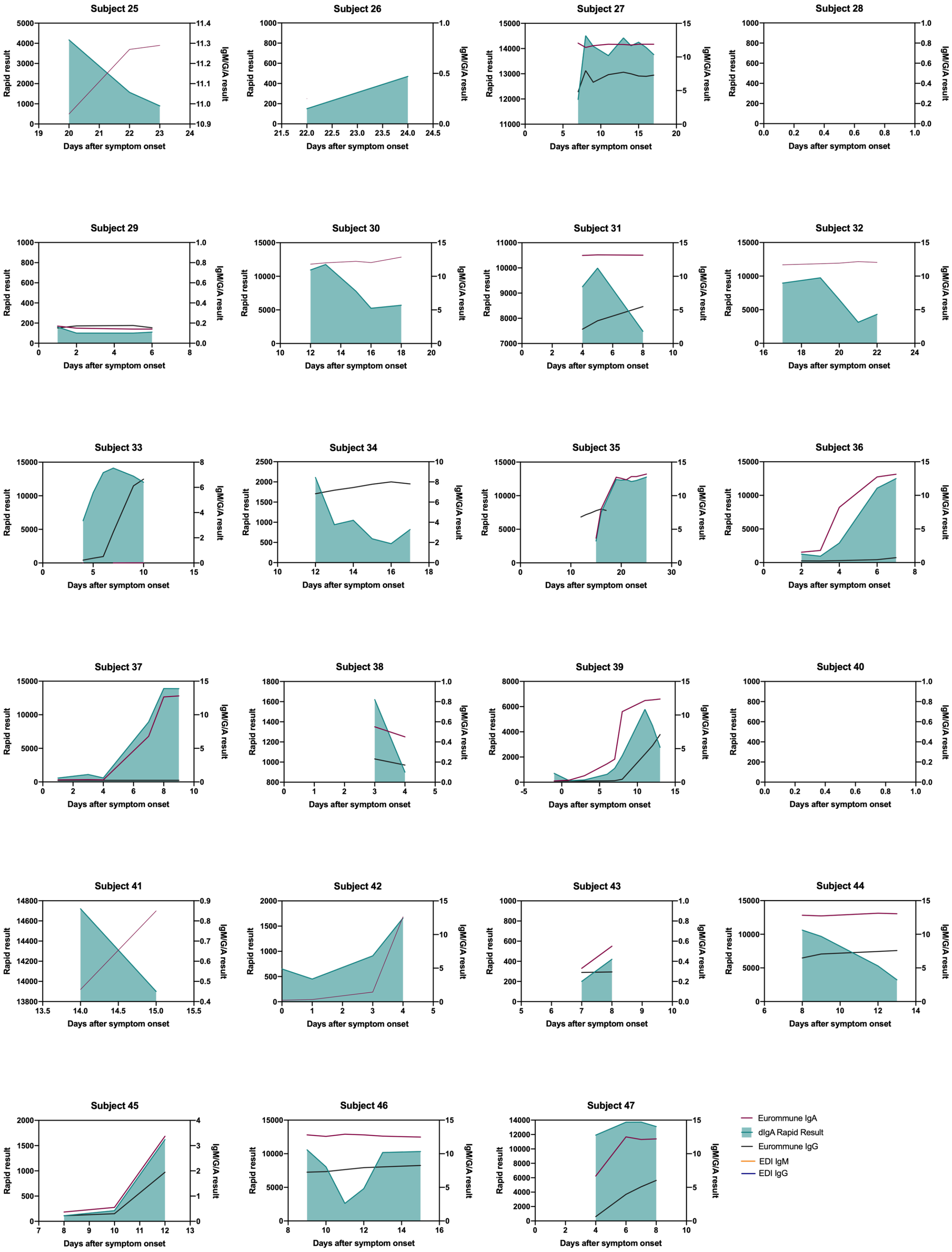


**Figure S5.** Subject by subject analysis of longitudinal dIgA, IgM, IgA and IgG responses towards SARS-CoV-2 antigens. Sera from available time points were analysed using the Rapid dIgA test, EDI IgG, EDI IgM and EUROIMMUN IgG and IgA test.

**Table S1.** Characteristics of the 22 non-SARS-CoV-2 coronavirus panel

| Sample ID | EDI-IgM Result | Rhinovirus/Enterovirus | Influenza | PCR result | | | | Parainfluenza | HIV | IgG result | | | | dIgA |
|---|---|---|---|---|---|---|---|---|---|---|---|---|---|---|
| | | | | 229E | HKU1 | NL63 | OC43 | | | 229E | HKU1 | NL63 | OC43 | |
| 86 | - | + | - | - | - | - | - | - | - | + | + | + | + | + |
| 85 | - | + | - | - | - | - | - | - | - | + | + | + | + | - |
| 69 | - | + | - | - | - | - | - | - | - | - | + | + | + | - |
| 68 | - | + | - | - | - | - | - | - | - | - | + | + | + | - |
| 67 | - | + | - | - | - | - | - | - | - | - | + | + | + | - |
| 93 | - | + | - | - | - | - | - | - | - | + | + | - | - | - |
| 73 | - | + | - | - | - | - | - | + | + | + | + | + | + | - |
| 82 | + | - | - | + | - | - | - | - | - | + | + | + | + | - |
| 44 | - | - | - | - | - | + | - | - | + | + | + | + | + | - |
| 95 | - | + | - | - | - | - | - | - | - | + | + | + | + | - |
| 84 | - | + | - | - | - | - | - | - | - | + | + | + | + | - |
| 83 | - | + | - | - | - | - | - | - | + | + | + | + | + | - |
| 57 | - | + | - | - | - | - | - | - | - | + | + | + | + | - |
| 60 | - | + | - | - | - | - | - | - | - | - | + | - | - | - |
| 100 | - | + | - | - | - | - | - | - | - | + | - | + | + | - |
| 58 | - | + | - | - | - | - | - | - | - | - | + | + | - | - |
| 80 | - | - | - | - | - | - | + | - | + | + | + | + | + | - |
| 81 | - | - | - | - | - | - | + | - | - | + | + | + | + | - |
| 55 | - | + | - | - | - | - | - | - | - | - | + | + | - | - |
| 91 | - | + | - | - | - | - | - | - | - | + | + | + | + | - |
| 61 | - | + | - | - | - | - | - | - | - | + | + | + | + | - |
| 98 | - | + | - | - | - | - | - | - | + | + | + | + | + | - |
